# Globalisation and Maternal Health Care Utilisation in sub-Saharan Africa: A Bayesian Multilevel Analysis

**DOI:** 10.1101/2022.06.02.22275939

**Authors:** Simona Simona

## Abstract

**Background:** Globalization is considered a major structural determinant of health and health care outcomes across the world. This study examines the influence of globalisation on maternal healthcare utilisation in sub-Saharan Africa.

**Methods:** Cross-sectional pooled survey data from 34 Demographic and Health Surveys (DHS) with 22559 randomly selected women of reproductive age group with a recent birth were combined with country-level globalisation indices from the KOF Swiss Economic Institute. Bayesian multilevel models were applied on maternal health care utilisation indicators (antenatal care visits, institutional delivery, and postnatal care check-ups) in relation to three dimensions of globalisation indices (economic, social, and political) and selected covariates.

**Results:** The results from the study demonstrate that the influence of globalisation on maternal healthcare depends on the dimension of globalisation. After controlling for selected individual and community-level factors, social globalisation is significantly associated with all the indicators of maternal healthcare utilisation while economic globalisation is significantly associated with antenatal care and postnatal check-ups for mothers and new-born babies.

**Conclusion:** More consideration in terms of public policy and further research, should be given to dimensions of globalisation which are more likely to influence maternal healthcare utilisation in sub-Saharan Africa.

## Introduction

The debate about the importance of macro-structures to population health outcomes has been ongoing for several decades now. Globalisation is included in this body of knowledge that attempts to regard health and healthcare outcomes as the function of not only biological, behavioural and lifestyle mechanisms but also of broader structural conditions that make up the social determinants of health ^1,2^. Globalization is a multifaceted term which renders itself to many definitions and controversies. Here we shall use Jenkins’s definition which regards globalization as “a process of greater integration within the world economy through movements of goods and services, capital, technology and (to a lesser extent) labour, which lead increasingly to economic decisions being influenced by global conditions” ^3^.

Globalization is often presented in global literature as having both negative and positive effects on health outcomes. Research in the world systems and dependency theory traditions for example, has probably been the most profound in highlighting the role that globalisation plays in exacerbating poor social, economic and health outcomes around the world. They mainly posit that unequal exchange relationships between the core and periphery countries result in the underdevelopment of the latter. It has been argued that economic linkages between developed and developing countries through trade, foreign direct investments and aid have been detrimental to health and quality of life in developing countries ^4–6^. The rationale behind this formulation is that globalisation increases the rate of economic growth but at the same time increases income inequality which adversely affects well-being for a large number of people especially in developing countries ^7,8^. Trade between the core and peripheral nations does not help this process because it is based on an unequal framework – the exchange of raw materials for processed goods and with long-term decline of prices for primary goods relative to finished goods. This results in weakened abilities to raise revenues by peripheral states and the lack of revenue limits the state’s health expenditure which would subsequently affect the funding of health programmes, recruitment and motivation of health personnel, provision of adequate health facilities and other social services that enhances life chances for many people ^7^. The quality of maternal health care is equally affected by lack of delivery care facilities and a cadre of lowly paid and demotivated staff.

However, some empirical studies have concluded that globalisation is good for public health, social development and developing countries’ food security ^9–13^. The mechanisms and pathways that make globalisation have a positive influence on population health are mainly economic in nature. Indeed, globalisation has been found to stimulate economic growth through increased employment opportunities, reduction of prices for consumer goods, encouragement of entrepreneurship and improved economies of scale in production among others ^14^. The precondition for this would be competitive domestic markets, strong regulatory institutions, moderate asset concentration and widespread access to public health services.

Globalisation can be linked with maternal health care utilisation through different pathways. The established association between globalisation and economic growth for example, implies improved education levels and employment status in the population. Studies in sub-Saharan Africa have shown that women who are educated, employed, or otherwise live with partners who are in the higher socioeconomic status bracket, are more likely to utilise maternal health care ^15–17^ Urbanisation is another intervening factor that has been found to link globalisation and population health ^18^. Since globalisation encourages urbanization in urban areas for instance, the distance to health facilities is not such a big problem. Also, that access to information about the dangers of delivering in homes and not visiting hospitals during pregnancy in urban areas. All these are drivers of positive maternal health care utilisation.

Many studies on the role of globalisation in health have often been criticised for their conceptualisation of globalisation as a solely economic process ^18^. There has been a move to a broader definition of globalisation that also captures the social and political aspects. Keohane and Nye ^19^, formulated a more widely accepted measurements of globalisation which includes three dimensions: a) economic: long distance flows of goods, capital, and services as well as information and perceptions that accompany market exchanges, b) political: the diffusion of government policies internationally, and c) social: the spread of ideas, information, images, and people. The KOF index of globalisation was created by Dreher and colleagues ^20^ to include the three and some more sub-dimensions. For all dimensions, the index was formulated using comprehensive data sets from 1970 to 2015.

Social and political globalisation have implications on the use of maternal health care in sub-Saharan Africa through the extent to which information is spread across the population and the impact of integration of global norms respectively. Social and cultural globalisation involves media freedom and cross-border movements of people and cultures. It is probable that women who have better exposed to technology, internet, mobile phones, and tourists are likely to value maternal health services and use them more than those who aren’t. Political globalisation on the other hand, has to do with participation in international norms and may also be a positive driver of maternal health care utilisation. Political globalisation is often regarded as a precursor to greater economic integration by encouraging trade between countries ^21^. Although the relationship between political globalisation and maternal health may not be straightforward, it is conceivable that those who live in countries that uphold international norms would use maternal health care services better than those who are not. This is what makes inclusion of the non-economic measures of globalisation important.

This study makes use of the new measurement and its different aspects to investigate the role of globalisation in maternal health care utilisation in sub-Saharan Africa. It is important to include all the different aspects of globalisation in the analysis because they capture different dimensions that may not be related to each other, making it possible to delineate which aspects may be responsible for women’s propensity to use maternal health care services. There are a few other studies that have used these measurements to study child mortality and obesity ^18,22^.

## Methods

### The data

The individual and community-level data (level 1 and 2) is derived from 34 Demographic and Health Surveys (DHS) conducted between 2006 and 2015 in sub-Saharan Africa. The Demographic and Health Surveys (DHS) are nationally representative population-based cross-sectional survey of women and men of reproductive age (15-49 year for women and 15-59 years for men) designed to provide information on a several measures, including maternal and child health. The DHS uses two-stage cluster sampling to arrive at eligible households based on the pre-existing sampling frame, typically the most recent census. Details of the DHS sampling procedure and sample sizes are explained elsewhere (Simona, 2020) and the data is available at the DHS program website: https://bit.ly/3NpP3UJ

The country-level data which forms the main explanatory variables are derived from the 2016 KOF globalisation index at the KOF Swiss Economic Institute. The globalisation indexes are from 1 up to 100 and the countries with higher scores are the most globalised ones and the ones with lower score are the least globalised. The definitions and measurements of the component variables of globalisation are described below and the data are available on the KOF Economic Institute website: https://bit.ly/3PqrcpB

### Variable selection and operationalisation

#### Outcome variables

The outcome variable for this study uses three dichotomous indicators of maternal healthcare utilisation from the DHS including antenatal care visits, institutional delivery and postnatal check-ups for mothers and new-born babies. Antenatal care visits mean the number of times women received antenatal care during their most recent pregnancy. The variable had two categories: ‘0’ = at least three times and ‘1’ = four or more times. Institutional delivery care was derived from a question of whether women delivered from; ‘0’ = home or ‘1’ institution. As for postnatal care, women were asked whether they had postnatal check-ups with their babies one month after delivery and their responses were also recoded into a binary variable taking the value of “0” if they were not checked and the value of “1” if they were checked.

#### Explanatory variables

Globalisation is the main explanatory variable which is delineated into four components including total, economic, social, and political globalisation. Total globalisation: This is an aggregation of the economic, social, and political components of globalisation. It is measured using the KOF Globalisation indicator.

Economic globalisation is a composite measure which is a component of the KOF globalisation index. It comprises trade in goods and services (as a percent of GDP), foreign direct investments (% of GDP), portfolio investments (% of GDP), international debt (% of GPD), international reserves and international income payments (% of GDP). Others are trade taxes and tariffs, capital account openness and investment restrictions. Social globalisation is based on the international voice traffic, transfers, international tourism foreign population, migration (% of total population), patent applications, international students, high technology exports, trade in cultural goods, McDonald restaurants (per capita), telephone subscriptions, international airports, television ownership, press freedom, internet, gender parity and expenditure on education.

Political globalisation is created from the number of embassies, participation in UN peace keeping missions, presence of international NGOs, international treaties signed and number of partners in investment treaties. This component is designed to measure the degree of a country’s international political engagement ^18,21^.

The study controls for important variables that may be associated with maternal health care utilisation or may be intervening in the relationship between globalisation and maternal health care localisation. These variables are defined at different the country, community, and individual levels and these include GDP per capita purchasing power parity at the country level. Media exposure, education attainment level, distance to health facility and place of residence are measured at the community level. The same variables are also measured at the individual level. Community-level factors were aggregates of individual level variables at the primary sampling units (PSU) or cluster level because the DHS only collects individual-level data. The aggregates were computed using the mean values of the proportions of women in each category of a given individual variable. Since the aggregate values may not have pragmatic meaning, the aggregate values of clusters were categorised into groups of ‘Lower’ and ‘Higher’ proportions based on national median values. Community education has three categories of ‘lower’, ‘middle’ and ‘higher’ while place of residence retains the original categorization of ‘rural’ and ‘urban’.

### Statistical analysis

Data extraction, management and analysis are all conducted the R statistical programming environment version 3.4.4 ^23^. There are several packages that have been used from preparation of data through to implementation of statistical modelling. Data manipulation and management relied mostly on the *dplyr* package ^24^. The analysis for the study started by mapping the distribution of the four components of globalisation across sub-Saharan African countries in 2015, followed by correlation analysis of the globalisation and the three indicators of maternal health care utilisation. Bayesian multilevel analysis was used to assess the influence of globalisation on maternal health care utilisation as well as examining relative country, community, and individual-level variations in maternal health care utilisation. Bayesian multilevel models were implemented using Markov chain Monte Carlo (McMC) methods in R2MLwiN ^25^.

Multilevel models were deemed appropriate for this analysis because it considers the nested structure of the data which is often difficult to represent in a single-level modelling approach. In this case, individual women are nested within communities, which are in turn nested within countries. Considering the hierarchical structure of the data set and the possible correlation that may exist within and between clusters and countries, a three-level random intercept logistic regression model was used.

I use uninformative uniform prior distribution for both regression and precision parameters in this study specifying 1 chain running for 55,000 iterations with a burn-in length of 5,000 iterations to achieve convergence. The convergence of chains was assessed by inspection of trace and auto-correlation plots. The Bayesian Deviance Information Criterion (DIC) was used to evaluate the goodness of fit of the models^26–28^.

For all the indicators of maternal healthcare utilisation, four models were specified for each of the outcome variables. In all the tables, model 1 is the null or empty model which contains only the intercept and the outcome variable. Model 2 includes all the three components of globalisation (economic, social, and political globalisation) which have been measured at the country-level. Model 3 includes community-level variables (community education, community distance to health facility problem, community media exposure and place of residence) and Model 4 controls for the individual level socio-demographic variables (educational status, distance to a health facility and exposure to the media). The tables report the posterior odds ratio (POR) and 95% Bayesian credible intervals (CrI) for each of the variables in all models except for the null where only the intercept parameters are reported. Statistical significance is determined by non-inclusion of “0” in the 95% (CrI).

I used the median odds ratios (MOR) and the variance partition coefficients (VPC) to examine the between country and between cluster variations in maternal health care utilization. The MOR is on the same scale as the odds ratios and is interpreted as the median value of the odds ratios between individuals from units at high or low risk when randomly choosing 2 individuals from different units. In this study, that would be the odds of having inadequate utilisation of maternal healthcare that are determined by unexplained factors at the community and country levels. The VPC provides information on the share of the variance at each level of analysis (individual, community, and country-levels). The VPC at each level was calculated using the latent method. It assumes a threshold model and approximating the level-1 (individual) variance by *π*^2^/3 (≈ 3.29) ^29–31^. Higher VPC values denote that a greater share of total variation in the outcome variables is attributable to higher level membership.

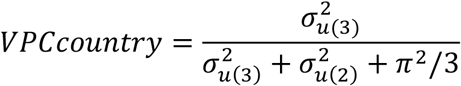

and

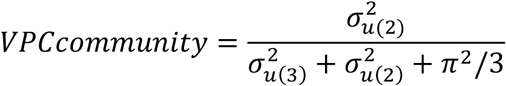

The Bayesian Deviance Information Criterion (DIC) was used to evaluate the goodness of fit of the models ^26–28^. When different models are compared, a smaller DIC means the model better fits the data than one with a high DIC value.

Ethical consideration

## Results

### Descriptive statistics

For this study, a total of 245955 women (level 1) were considered, nested within 17,871 clusters (level 2), within 34 sub-Saharan African countries (level 3). Suppl Table 1. presents the 34 sub-Saharan African countries that are included as well as survey years, final sample per country, number of communities in a country, median number of respondents per community and range of respondents in a community. The distribution of maternal health care utilisation across sSA countries is shown in the Suppl Fig.1. Figure 1. shows the distribution of the three components of globalisation indexes and the aggregated total globalisation across sSA countries in 2015. It is interesting to note that the performance of countries varies in each of the globalisation indexes. Liberia, Namibia, and Nigeria are the most globalised in terms of economic, social, and political globalisation respectively. In terms of aggregated globalisation however, it is evident that Senegal, Namibia, Ghana, and Kenya are among the most globalised countries while countries like Burundi, Chad, Comoros, and Congo DR are some of the least globalised in the subcontinent.

**Figure 1.**
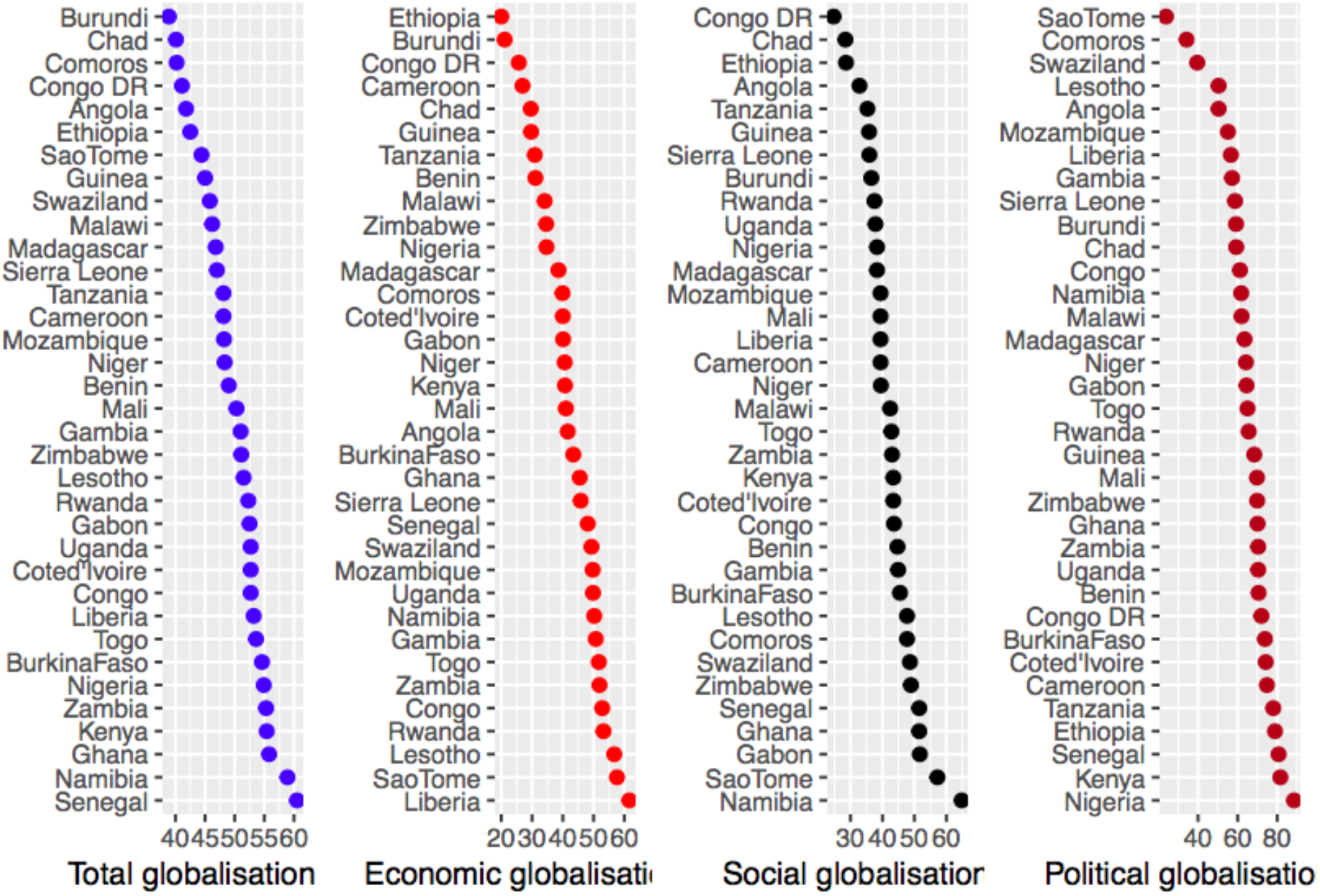
The distribution of the four components of globalisation across sub-Saharan African countries

### Correlation between globalisation Indexes and Maternal healthcare indicators in

Figure 2. displays results of non-parametric associations between the four components of globalisation and the proportion of having 4 or more antenatal care visits in sSA. This is a pooled analysis of correlation coefficients of each of the four components of globalisation and the proportion of having adequate antenatal care visits among women in selected sSA countries. All countries selected in the analysis are included except for overlapping ones which were removed to ensure quality visualisation. Correlation coefficients are reported and indicated on top of each graph with the linear line of fit and 95% confidence interval indicated by the shaded areas. The results here indicate temporal associations between components of globalisation and maternal healthcare utilisation in sSA.

**Figure 2:**
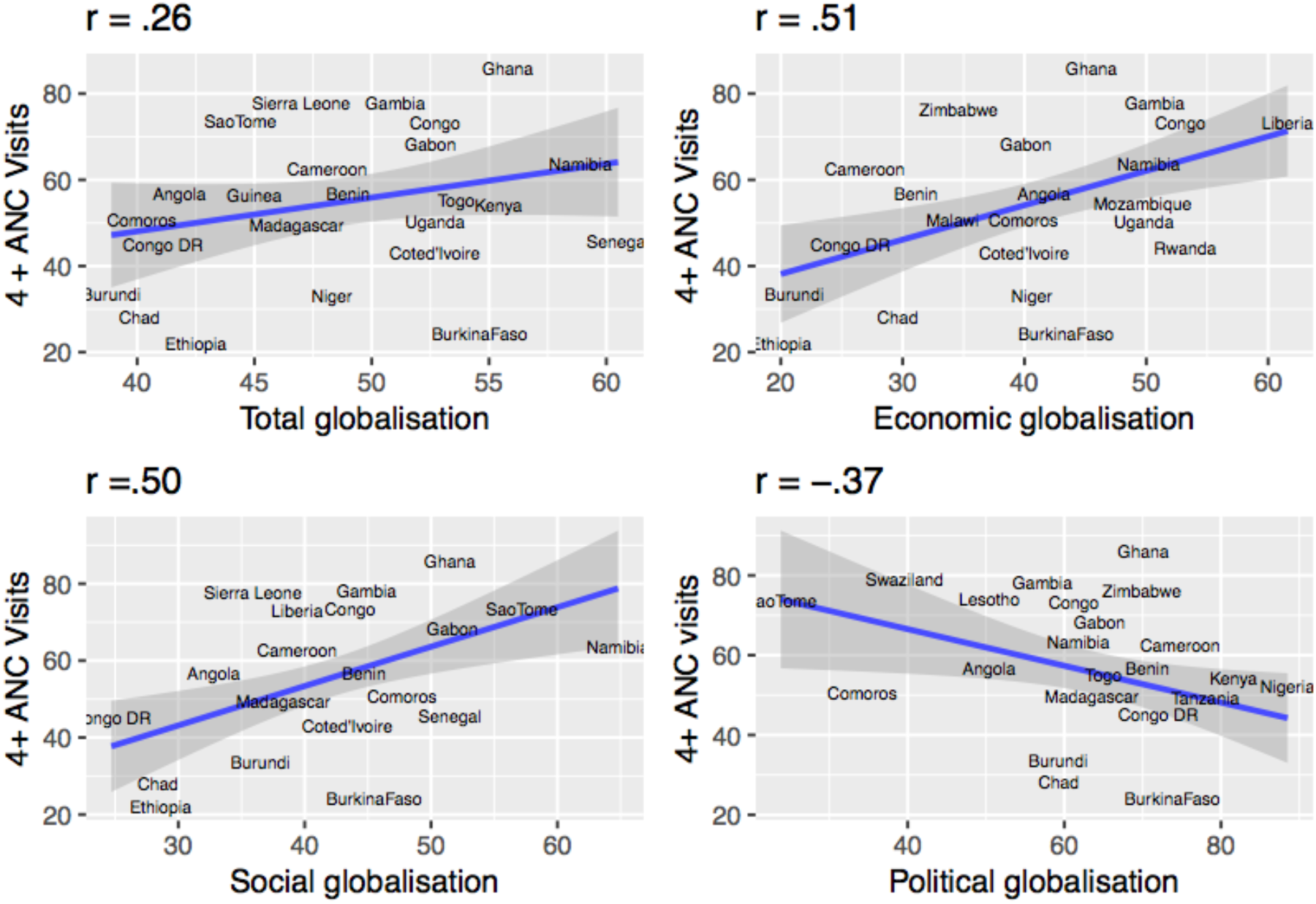
The relationship between the four components of globalisation and antenatal care visits

Apart from political globalisation, all the components of globalisation show moderate but positive correlation with the proportion of adequate antenatal care visits. Total globalisation is not significantly related to antenatal care while economic and social globalisation display significant relationships (t = 3.34) and (t = 3.45) respectively. Political globalisation on the other hand, shows a negative and significant relationship with the proportion of 4 + more visits (t = -2.30).

In Fig. 3. I plot the correlation between the four components of globalisation and the proportion of institutional delivery in sSA. There is a linear, positive, and moderate correlation between 3 components of globalisation (total, economic and social globalisation) but the proportion of institutional delivery is only strongly correlated with total and economic globalisation. Just like in the case of antenatal care visits, political globalisation is negatively correlated with institutional delivery.

**Figure 3:**
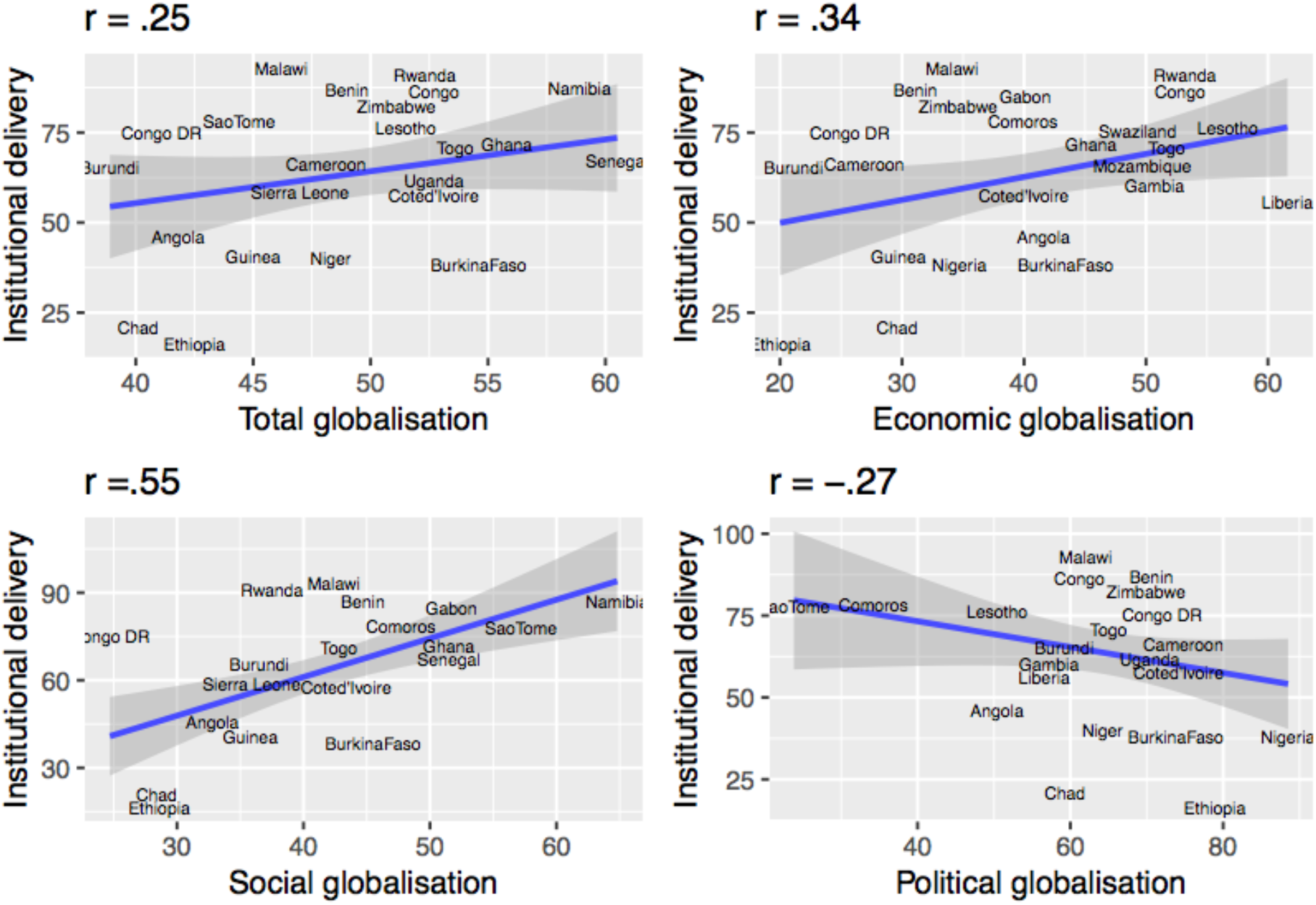
The relationship between the four components of globalisation and institutional delivery

**Figure 4.**
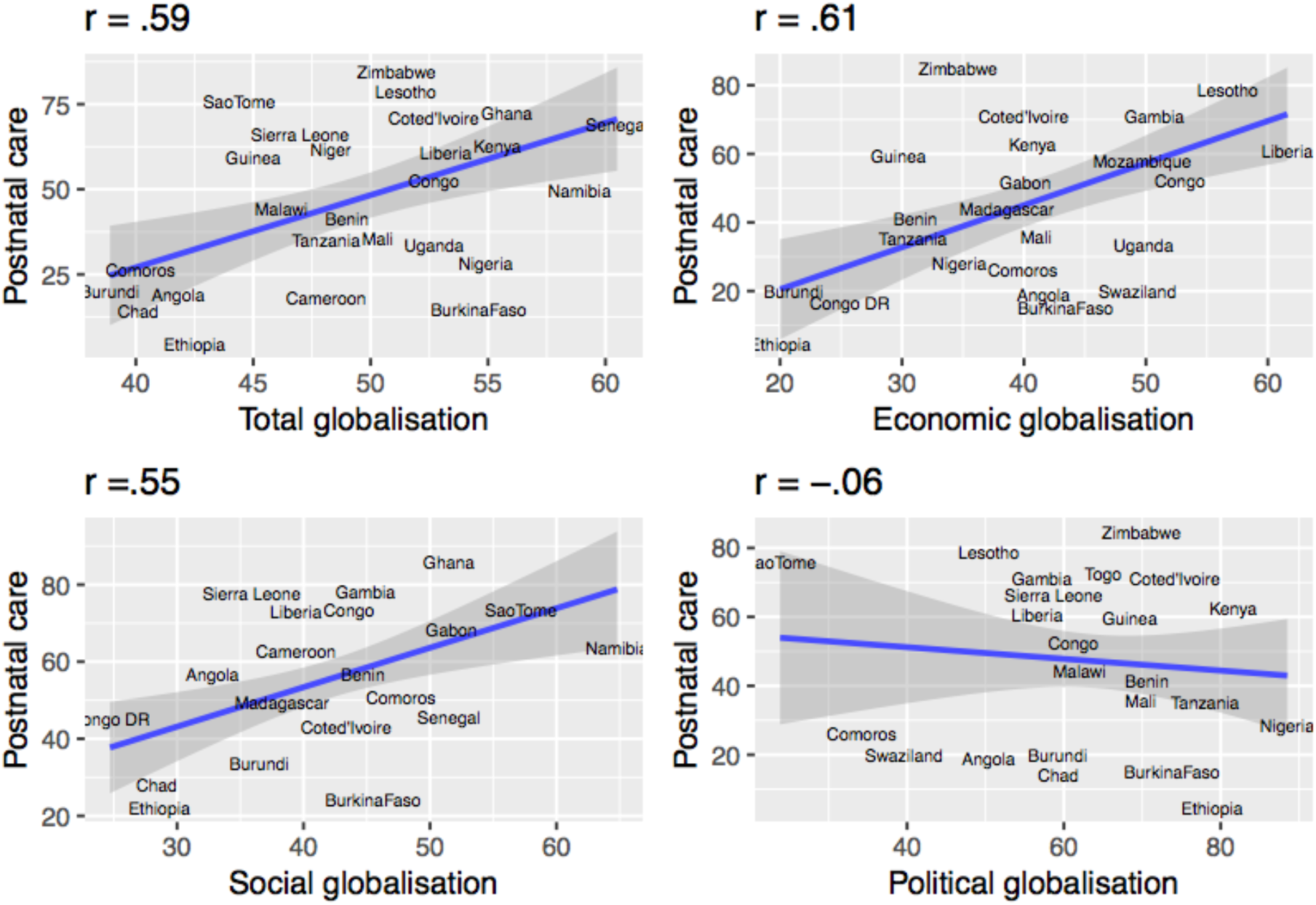
The relationship between the four components of globalisation and postnatal care

The relationship between the four components of globalisation and postnatal care shows a similar pattern as the other outcome variables except that in this case the 3 components are not only positively correlated with postnatal care, but the correlation is also statistically significance. Political globalisation still posits negative correlation albeit marginal.

### Multilevel analysis of the influence of globalisation on maternal healthcare utilisation

The non-parametric analysis undertaken above has a weakness of not indicating the extent of association between variables, neither does it enable prediction of the value of the outcome variable based on the explanatory variables. Even more importantly, issues of dependence, contextuality and heterogeneity in data are not captured in correlation analysis. As such, this part of analysis uses multilevel models to study the influence of globalisation on maternal health care utilisation. Fixed and random effects of each component of globalisation is modelled on each maternal health care indicator and presented in Tables 1-3. The Tables report the relationship between the three components of globalisation (economic, political, and social) and indicators of maternal healthcare utilisation, reporting posterior odds ratio (POR) and 95% Bayesian credible intervals (CrI) for each of the variables in all models except for the null where only the intercept parameters are reported. Statistical significance is determined by non-inclusion of “1” in the 95% (CrI). All significant results are highted. Total globalisation index being an aggregation of the three components is removed from the multilevel regression analysis due to multicollinearity.

**Table 1.**
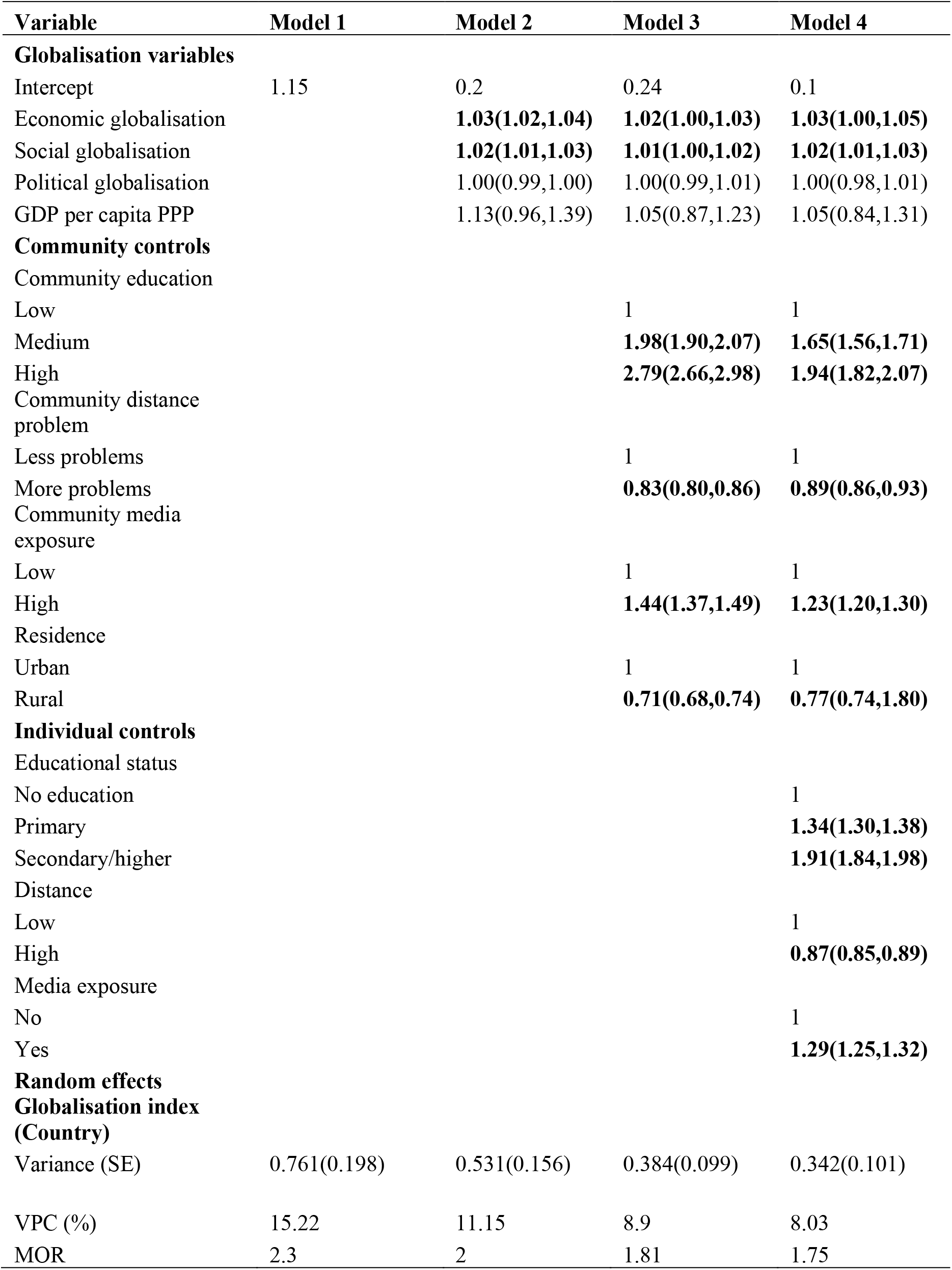
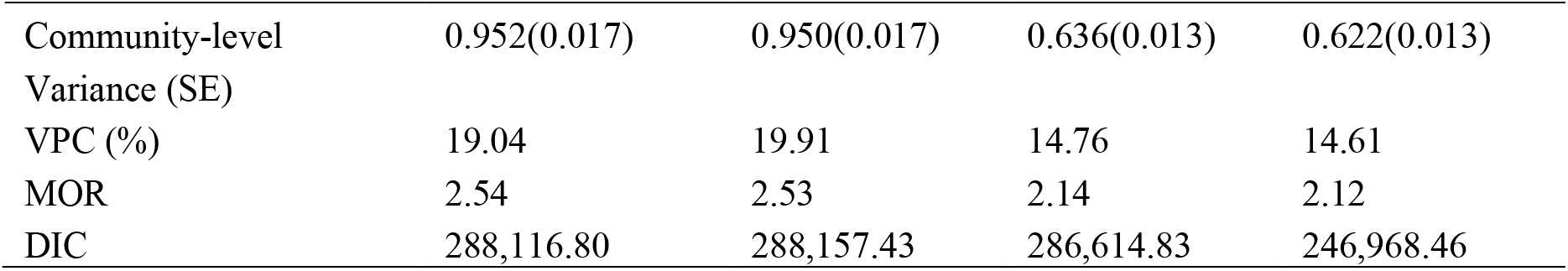
Posterior odds ratios for multilevel logistic regression for globalisation and antenatal care in sub-Saharan Africa with 95% credible intervals (N = 245955)

| Variable | Model 1 | Model 2 | Model 3 | Model 4 |
| --- | --- | --- | --- | --- |
| **Globalisation variables** |  |  |  |  |
| Intercept | 1.15 | 0.2 | 0.24 | 0.1 |
| Economic globalisation |  | **1.03(1.02,1.04)** | **1.02(1.00,1.03)** | **1.03(1.00,1.05)** |
| Social globalisation |  | **1.02(1.01,1.03)** | **1.01(1.00,1.02)** | **1.02(1.01,1.03)** |
| Political globalisation |  | 1.00(0.99,1.00) | 1.00(0.99,1.01) | 1.00(0.98,1.01) |
| GDP per capita PPP |  | 1.13(0.96,1.39) | 1.05(0.87,1.23) | 1.05(0.84,1.31) |
| **Community controls** |  |  |  |  |
| Community education |  |  |  |  |
| Low |  |  | 1 | 1 |
| Medium |  |  | **1.98(1.90,2.07)** | **1.65(1.56,1.71)** |
| High |  |  | **2.79(2.66,2.98)** | **1.94(1.82,2.07)** |
| Community distance problem |  |  |  |  |
| Less problems |  |  | 1 | 1 |
| More problems |  |  | **0.83(0.80,0.86)** | **0.89(0.86,0.93)** |
| Community media exposure |  |  |  |  |
| Low |  |  | 1 | 1 |
| High |  |  | **1.44(1.37,1.49)** | **1.23(1.20,1.30)** |
| Residence |  |  |  |  |
| Urban |  |  | 1 | 1 |
| Rural |  |  | **0.71(0.68,0.74)** | **0.77(0.74,1.80)** |
| **Individual controls** |  |  |  |  |
| Educational status |  |  |  |  |
| No education |  |  |  | 1 |
| Primary |  |  |  | **1.34(1.30,1.38)** |
| Secondary/higher |  |  |  | **1.91(1.84,1.98)** |
| Distance |  |  |  |  |
| Low |  |  |  | 1 |
| High |  |  |  | **0.87(0.85,0.89)** |
| Media exposure |  |  |  |  |
| No |  |  |  | 1 |
| Yes |  |  |  | **1.29(1.25,1.32)** |
| **Random effects** |  |  |  |  |
| **Globalisation index (Country)** |  |  |  |  |
| Variance (SE) | 0.761(0.198) | 0.531(0.156) | 0.384(0.099) | 0.342(0.101) |
| VPC (%) | 15.22 | 11.15 | 8.9 | 8.03 |
| MOR | 2.3 | 2 | 1.81 | 1.75 |
| Community-level | 0.952(0.017) | 0.950(0.017) | 0.636(0.013) | 0.622(0.013) |
| Variance (SE) |  |  |  |  |
| VPC (%) | 19.04 | 19.91 | 14.76 | 14.61 |
| MOR | 2.54 | 2.53 | 2.14 | 2.12 |
| DIC | 288,116.80 | 288,157.43 | 286,614.83 | 246,968.46 |

### Globalisation and antenatal care in sSA

The results of the influence of globalisation on antenatal care visits in sSA are reported in Table 1. Since this study delineates the concept of globalisation into its distinct components to determine whether the components indeed capture different phenomena, each of the components were regressed on antenatal care visits. Results from fixed effects here show that economic and social globalisations are significantly associated with antenatal care visits albeit with small effect sizes. This means that women who live in countries with higher economic and social globalisation have increased odds of having adequate number of antenatal care visits during pregnancy. A unit increase in economic globalisation index increases the odds of four or more antenatal care visits by a factor of 1.03 (95% CrI 1.00-1.05) after controlling for the effects of covariates. After community and individual level variables are controlled for, a unit increase in globalisation increases the odds of having four or more antenatal care visits by a factor of 1.02 (95% CrI 1.01-1.03).

Political globalisation and GDP per capita PPP are not significantly associated with antenatal care visits in sSA. Control variables at the community and individual level are found to be significantly associated with antenatal care. For example, living in communities where most women are higher (OR = 1.94, 95% CrI 1.82-2.07) or moderate education (OR = 1.65, CrI 1.56-1.71), with exposure to the media (OR = 1.23, 95% CrI 1.20-1.30) increases the odds of adequate antenatal care visits, while living in rural areas (OR = 0.77, 95% CrI 0.74-1.80) with longer distances to healthcare facilities (OR = 0.89, 95% CrI 0.86-0.93) decreases the odds of having four or more antenatal care visits during pregnancy. All the individual level covariates were associated with antenatal care.

Model 1 shows significant variations in the odds of antenatal care visits across 34 sub-Saharan African countries (*σ*^2^ = 0.76) and across communities (*σ*^2^ = 0.95) respectively. The VPC results indicate that 15.2% of cross-national variations in antenatal care is attributed to the country-level factors while 19% is attributable to the community level characteristics. Although, the greatest contribution to variations in antenatal care remains at the individual level, community and country level factors contribute significantly to explaining cross-country variations in sSA. The reduction in the value of VPC after the introduction of country, community and individual level factors into the models equally indicates the importance of considering community and country level factors. Results from the MOR also helps to confirm evidence of community and country characteristics shaping antenatal care visits. From Model 4, it was estimated that if a women moved to another country or community with a higher probability of antenatal care visits during pregnancy, the median increase in their odds of having four or more visits would be 1.75 and 2.12 respectively. Compared to the MOR of one which is the threshold of no variance, these estimates show significant community and country variances in antenatal care visits.

Table 1 also shows that the full model (including the country, community, and individual level factors) has lower computed BIC values than the proceeding models. This means that the full model fits well to identify the factors influencing antenatal care visits in sub-Saharan Africa.

### Globalisation and institutional delivery in sSA

In Table 2, only social globalisation is significantly associated with antenatal care visits in sSA after all the country, community and individual level factors are controlled for. The odds of delivering in an institution are significantly higher in countries with higher social globalisation. This indicates that for one standard deviation increase in social globalisation index increases the odds of institutional delivery by a factor of 1.07 (95% CrI 1.07–1.08). Economic globalisation, political globalisation and GDP were all not associated with institutional delivery.

**Table 2.**
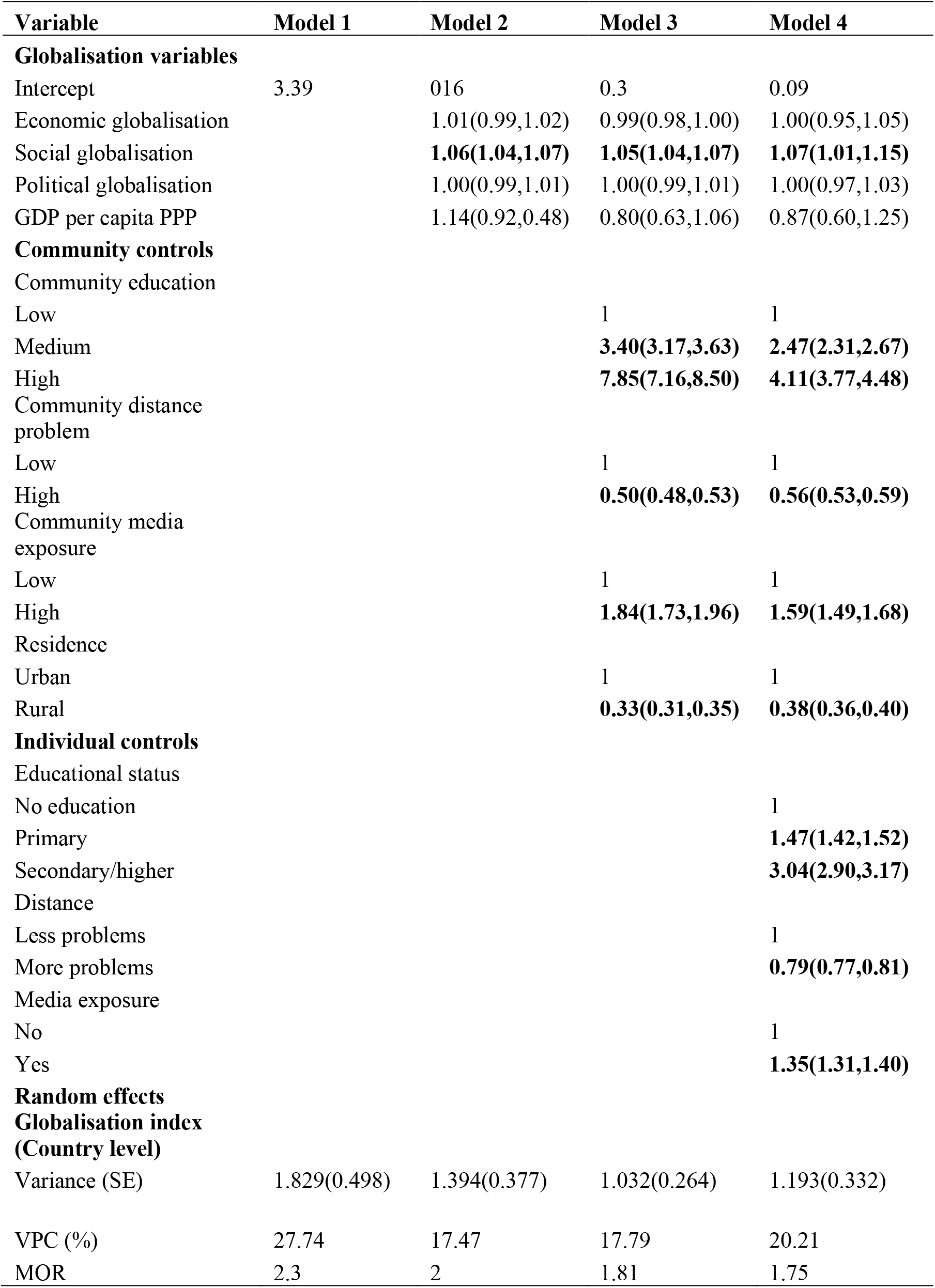
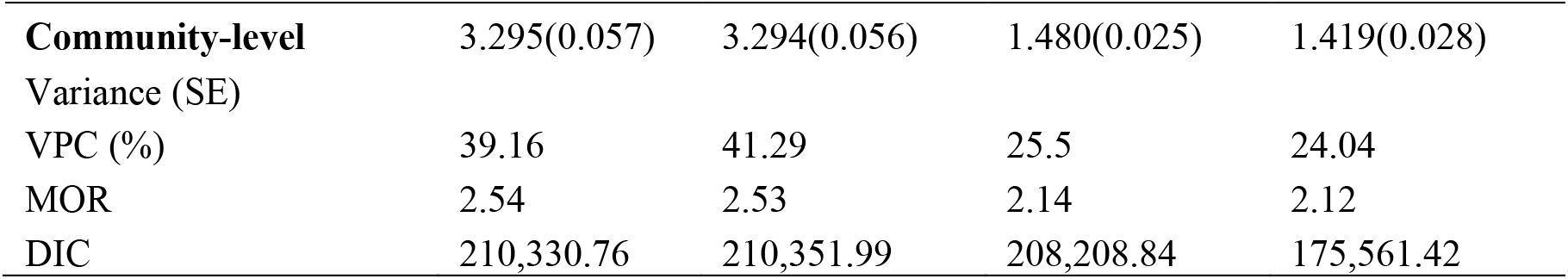
Posterior odds ratios for multilevel logistic regression for Globalisation and Institutional delivery care in sub-Saharan Africa with 95% credible intervals (N = 245955)

All the community and individual level controlling factors were significantly associated with institutional delivery. The results revealed that living in communities where most women have higher (OR = 4.11, CrI 3.77-4.48) or medium education (OR = 2.47, CrI 2.31-2.67) as well as exposed to the media (OR = 1.59, CrI 1.49-1.68) increases the odds of institutional delivery. On the other hand, rural residence (OR = 0.38, 95% CrI 0.36-0.40) and longer distances to health facilities (OR = 0.56, 95% CrI 0.53-0.59) reduces the odds of institutional delivery.

Significant between country (*σ*^2^ = 1.83) and between community (*σ*^2^ = 3.30) variations in institutional delivery were found and are reported in Model 1. Accordingly, the VPC reported indicates that the combined contribution of community and country-level factors in cross-national variation in institutional delivery is higher than individual level factors. Country-level and community level factors account for 27.7% and 39.2% variations in institutional delivery respectively. The MOR, reported to be 2.3 at the country level and 2.5 at the community level shows the importance of context in institutional delivery. The significant drop in the VPC at country-level from 27.7% to 17.5% after the community level factors are introduced shows the established importance of community-level factors in determining health and well-being. The decreasing values of the DIC with additional variables indicates that the full model is better compared to preceding models in explaining variations institutional delivery.

### Globalisation and postnatal care in sSA

Table 3 reports the results on the influence of globalisation on postnatal check-ups for mothers and new-born babies in sSA. All components of globalisation are significantly associated with postnatal care check-ups in before covariates are accounted for. However, after controlling for the effects of country, community and individual level factors, political globalisation lost significance. The results indicate that for a unit standard deviation increase in economic globalisation index, the odds of postnatal care check-ups increase by a factor of 1.06 (95% CrI 1.02-1.10), while a unit standard deviation increase in social globalisation increases the odds of postnatal check-ups by a factor of 1.08 (95% CrI 1.03-1.14).

**Table 3.**
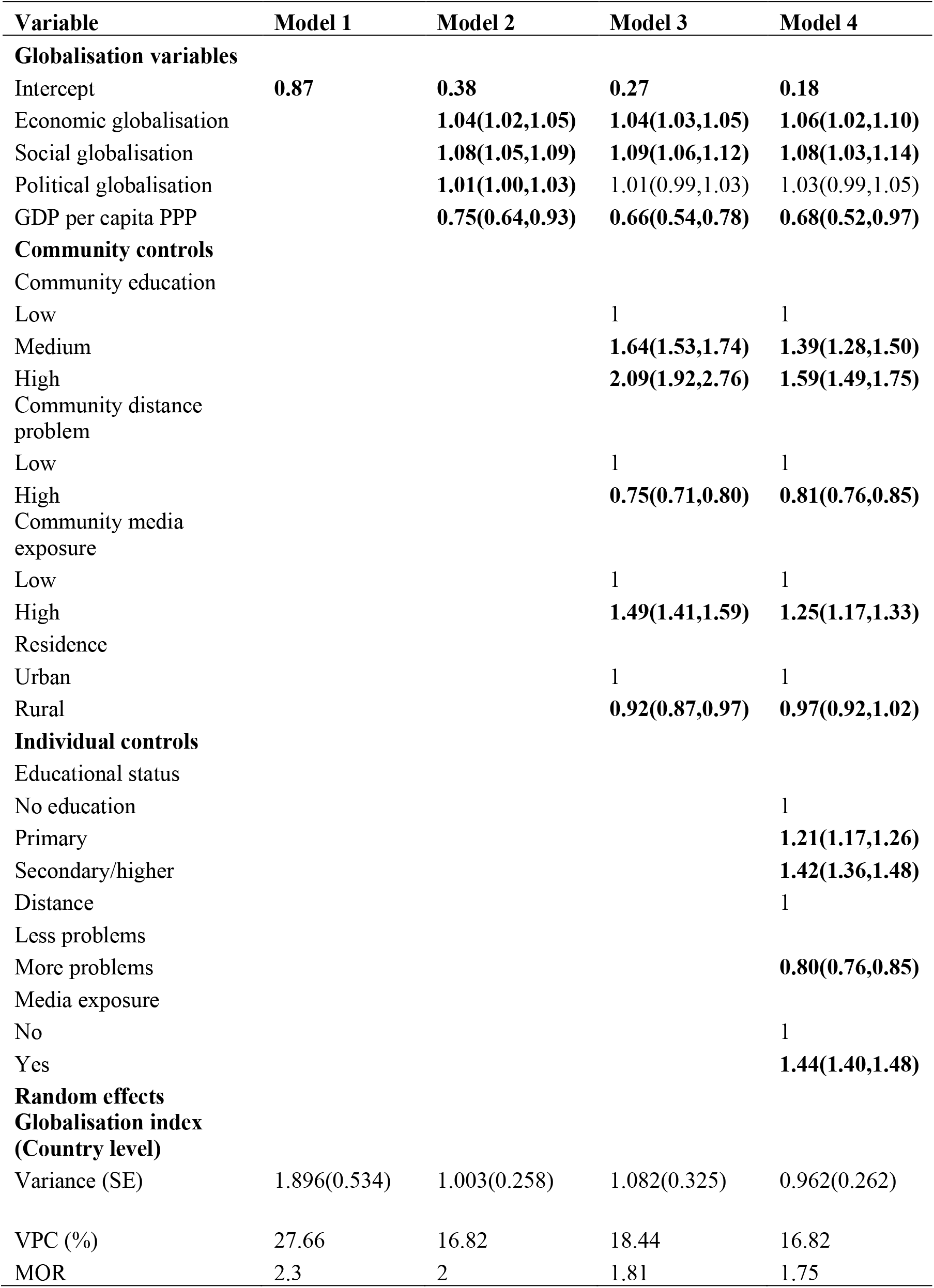
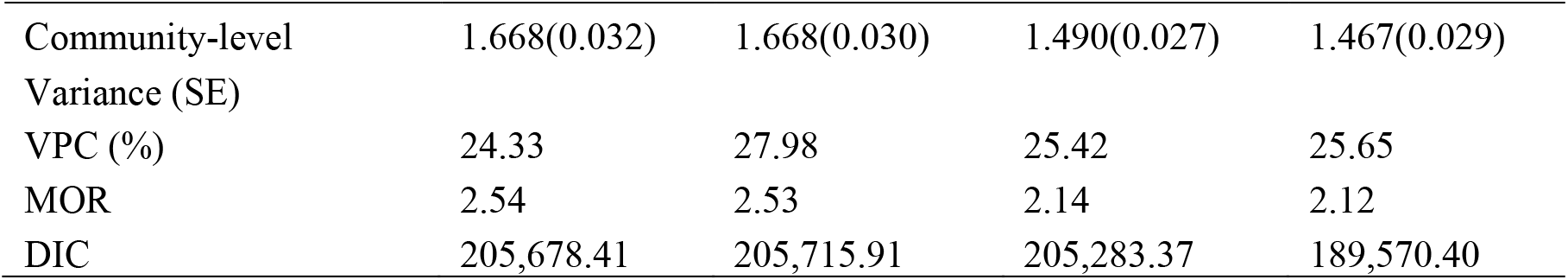
Posterior odds ratios for multilevel logistic regression for Globalisation and Postnatal care in sub-Saharan Africa with 95% credible intervals (N = 245955)

The fact that political globalisation lost significance after controlling for covariates means that some of the community and individual level variables share in the explanatory power. The results also show surprising finding in that GDP per capita PPP was negatively associated with postnatal care in sSA. Community characteristics which are significantly associated with postnatal care include being in communities with high proportion of women with high (OR = 1.59, CrI 1.49-1.75) or moderate education (OR = 1.39, CrI 1.28-1.50) as well as being from communities with most women exposed to the media (OR = 1.25, CrI 1.17-1.33). Also, women from communities where most women have problems accessing health facilities especially in rural areas have reduced odds of having postnatal check-ups.

Like in other indicators of maternal healthcare utilisation, significant between country (*σ*^2^ = 1.90) and between community (*σ*^2^ = 1.76) variations are also reported in Model 1. The VPC confirms the importance of country (28%) and community level (24%) in explaining variations in postnatal care check-ups among mothers and new-born babies. Furthermore, greater deviation of the MOR values from one at the country (2.30) and community (2.54) levels also indicate the importance of contextual variables in understanding cross-national variations in postnatal care. Like in the other models for antenatal care and institutional delivery, the country-level VPC drops significantly when country-level and community-level factors are introduced into the models. This, as already stated further signifies the importance of contextual level factors in explaining maternal health care in sSA.

## Discussion

The study examined the influence of globalisation on the maternal health care utilisation continuum of antenatal care visits, institutional delivery and postnatal check-ups for mothers and new-born babies in sSA. The concept of globalisation was delineated into three dimensions of economic, social, and political, to distinguish the influence of many different processes that made up globalisation. The results from this study support the notion that the influence of globalisation on health depends on particular dimensions studied^11,18,32^. Results from fixed effects showed that social globalisation is the most consistent predictor of maternal healthcare across the continuum of antenatal, institutional delivery and postnatal care. Economic globalisation is significantly associated with antenatal care and postanal check-ups for mothers and new-born babies. In adjusted models, political globalisation was found to have no significant influence on any indicators of maternal healthcare utilization in sub-Saharan Africa.

Social globalisation encompasses the domain of information sharing, exchange of ideas as well as the technology and freedom to do so. It is plausible that women who live in countries with high social globalisation index would be more likely to use maternal healthcare compared to their counterparts who do not. Several studies have demonstrated the crucial role played by exposure to the media as well as civil and socio-economic rights in maternal and reproductive health matters ^33–35^. Promotion of civil, media, technology, and communication freedoms shape culture, enhance citizen awareness about their rights, and influence national politics which may in turn lead to better provision, access, and utilisation of health services. Social globalisation and political globalisation were found to be a consistent predictor of overweight and obesity in a study that focussed on low and middle income countries^18^. The current study found no significant relationship between political globalisation and maternal healthcare utilisation.

Economic globalisation was only found to have a positive influence on antenatal and postnatal care and the analysis of these relationships include a more traditional global narrative which considers globalisation as a precursor of global economic growth funnelled through weakened relevance of global boundaries, greater interdependence between countries and increased adherence to international norms ^36,37^. Weakened borders and interdependence between nations support trade liberalism and other economic processes such as foreign direct investments, which in turn leads to economic growth and associated population health and well-being ^21^. Adherence to international norms means greater propensity of implementing international treaties which may border on human rights obligations which require states to deliver socio-economic entitlements including health and health care ^37^.

This study accounted for heterogeneity across countries and communities included. Using three-level Bayesian models, contextual factors were found to have significant influence on maternal healthcare utilisation. This finding may not only imply that country and community characteristics are directly associated with indicators of the maternal healthcare continuum but also that variations may come from individual or compositional effects with particular types of people who are more likely to have lower odds of maternal healthcare utilisation on account of their individual characteristics being found more commonly in certain communities and countries^38^. In this regard, it is evident that living in countries with high globalisation index, especially social and economic as well as living in urban areas, with shorter distances to health facilities and high proportions of educated women, matters with respect to maternal healthcare utilisation.

### Strengths and limitations of the study

Studying the influence of globalisation on maternal health care using Bayesian three-level multilevel models is novel. The study has established not only the extent of the influence of globalisation on maternal healthcare utilisation but also that overall, contextual factors account for more variations in institutional delivery and postnatal care than individual factors. These findings provide evidence on the importance of broader structural determinants of health and well-being in sub-Saharan Africa. The study also offers an important contribution in the empirical conceptualisation of globalisation, considering it as a composite term including different dimensions such as economic, social, and political. To my knowledge there is no previous study which focusses on globalisation and maternal health care which using the three constituent dimensions.

Inevitably this study has limitation just like any other that uses the DHS or any cross-sectional survey. The analysis is only able to determine associations or relationships between variables and not cause and effect. It is also important to note that while study that determine causation will be desirable, the data infrastructure in sub-Saharan Africa is not well-developed to obtain data that will enable causality studies. Additionally, the study analysed data on women of the reproductive age group who have had birth within five years prior to the most recent DHS in included countries. Although this is not necessarily a limitation of the study as it is the function of the study design, it is important that the interpretation of the results is restricted to a specific section of the women population and not the entire population. Furthermore, countries that form high-level units in the multilevel analysis are relatively few and non-sample, which may generate imprecise parameter estimates. However, this limitation is counteracted by the use of Bayesian inference and Markov chain Monte Carlo estimates which have often been found to be more reliable even if the high-level units are relatively fewer ^39,40^.

## Conclusion

This study brings an important dimension to the study of maternal healthcare in sSA by identifying social and economic globalisation as determinants. The fact that social and economic globalisation were significantly associated with maternal healthcare utilisation even with relatively fewer number of countries is an indication that the relationship may be much stronger with a larger sample size. The implication from this study is therefore, that further research is warranted to not only validate the relationship between globalisation and maternal healthcare using bigger sample sizes, but also to understand the specific causal pathways between the two variables. I found for example that social globalisation is associated with all the three indicators of maternal healthcare utilisation, but social globalisation is itself a composite variable consisting of several components. It would be interesting to isolate and assess the influence of each element of social globalisation such as international tourism, migration, high technology exports, McDonald restaurants (per capita), international airports, television ownership, press freedom, gender parity and expenditure on education on maternal healthcare utilisation in sSA.

## Data Availability

The data used in this study and publicly available and no further ethical approval was needed

https://bit.ly/3PqrcpB

https://bit.ly/3NpP3UJ

## Author contribution

### Simona Simona

Conceptualization; Formal analysis; Investigation; Methodology; Software; Visualization; Writing— original draft; Writing—review & editing.

## Ethical approval and consent to participate

No further ethical approval was needed for this study because it used secondary data which is publicly available. The respondents did not participate in the study directly.

## Availability of data and materials

The data used in this study is a combination of globalisation indices from the KOF Swiss Economic Institute available: https://bit.ly/3PqrcpB and the DHS data which are available here: https://bit.ly/3NpP3UJ

